# A within-host model of SARS-CoV-2 infection

**DOI:** 10.1101/2022.04.22.22274137

**Authors:** Jonathan Carruthers, Jingsi Xu, Thomas Finnie, Ian Hall

## Abstract

Within-host models have been used to successfully describe the dynamics of multiple viral infections, however, the dynamics of SARS-CoV-2 virus infection remain poorly understood. A greater understanding of how the virus interacts with the host can contribute to more realistic epidemiological models and help evaluate the effect of antiviral therapies and vaccines. Here, we present a within-host model to describe SARS-CoV-2 viral dynamics in the upper respiratory tract of individuals enrolled in the UK COVID-19 Human Challenge Study. Using this model, we investigate the viral dynamics and provide timescales of infection that independently verify key epidemiological parameters important in the management of an epidemic. In particular, we estimate that an infected individual is first capable of transmitting the virus after approximately 2.1 days, remains infectious for a further 8.3 days, but can continue to test positive using a PCR test for up to 27 days.

## Introduction

The SARS-CoV-2 virus has led to a global pandemic, causing millions of infections world-wide. Mathematical modelling has been used throughout the pandemic to predict numbers of cases, hospitalisations and deaths, as well as the impact of non-pharmaceutical interventions [1, 2, 3]. In these epidemiological models, the rate at which susceptible individuals become infected can be thought of as the product of a contact rate and a transmission probability. The contact rate is simply interpreted as the number of contacts an individual makes per unit time, with insights provided from social contact studies [4, 5]. The probability of transmission, however, is not as well-defined and depends on many factors such as the environment in which the contact takes place, the duration of the contact and the infectiousness of the individual. Intuitively, an individual is most infectious when viral loads in their respiratory tract peak [6, 7], therefore understanding how viral loads change over time is crucial to understanding infectiousness. Within-host models can be used to describe how interactions between virus particles, host cells and the immune system affect the viral load over time, with the link to infectiousness then provided by either dose-response relationships or the assumption that the transmission probability is an increasing function of viral load [8, 9].

Existing within-host models of SARS-CoV-2 have predominantly focused on describing the dynamics of the virus in the upper respiratory tract (URT), the primary site of infection, where data are readily available from nasopharyngeal swabs [10, 11, 12]. In some cases, these models are extended to include compartments such as the lower respiratory tract (LRT), however, observing LRT infection is difficult and is unlikely to play a role in person-to-person transmission [13]. Often, swab data are presented in the form of polymerase chain reaction (PCR) cycle threshold values (Ct values) that are converted into RNA copies by means of a standard curve. However, since a PCR test captures all genetic material, it quantifies the total viral RNA and not the amount of viable virus that is capable of infecting cells [14]. This therefore makes it difficult for models parameterised using such data to predict the duration that an individual remains infectious; a previous study has shown that live virus cannot be isolated from respiratory samples 8-9 days after symptom onset notwithstanding persistently high viral RNA loads [15]. Despite this, some models have directly estimated the infectiousness of an individual from predictions of total viral load [16, 17], whilst others have first linked total and infectious viral loads using the probability that a sample is culture positive [18].

A second limitation of earlier within-host models is that swab samples were understandably only collected once an individual first displayed symptoms, so the exact date of infection is unknown. The majority of models account for this by assuming a fixed incubation period of between 3 and 7 days, however, since symptom onset approximately coincides with peak viral loads, the models can only reliably predict the decay in viral loads and not the initial growth [19]. Human challenge studies allow us to study the processes of infection and immunity from their inception and therefore provide a complete overview of the disease course [20]. Screening of participants can also ensure that samples are representative of primary infection, whilst a quarantine setting allows data to be obtained with a level of detail not achievable in a real-life setting [21].

In this paper, we develop a mathematical model to describe the dynamics of wild-type SARS-CoV-2 infection in the URT. We make use of infectious and total virus time courses from participants of a human challenge study, thereby addressing the above limitations of existing models. Bayesian inference methods are applied to estimate model parameters and the within-host basic reproduction number. The mathematical model is then used to predict timescales of infection, such as the duration that an infected individual remains infectious and PCR positive.

## Methods

### Model structure

The viral dynamics in the mid-turbinates of human challenge study participants are described using a target-cell limited model that has successfully been used to describe the dynamics of other viral infections, and makes use of measurements of both infectious and total viral loads [22, 23, 24]. In this model, target cells (*T*) become infected with infectious virus (*V*_inf_) at rate *β* and enter a non-productive eclipse phase (*E*) for 1*/k* days, after which they enter a productively infectious phase lasting for a further 1*/δ* days. Whilst cells are in the infectious phase, they produce infectious virus at rate *p*_inf_ which in turn loses infectivity at rate *c*_inf_. Viral RNA is produced by an infectious cell at rate *p*_tot_ and lose viability with rate *c*_tot_. To represent an adaptive immune response, we note that effector cell responses (*C*), such as those of CD8^+^ T cells, go through an antigen-dependent activation stage, followed by a virus-independent expansion phase. The growth rate of the effector cell population is therefore initially dependent on the total viral load (*V*_tot_), but once this becomes large enough, exponential growth of the effector cell population follows [25]. Effector cells help to resolve infection through the mass action killing of infected cells with rate *k*_*C*_ and survive for an average of 1*/δ*_*C*_ days. The model is described by the system of ordinary differential equations:

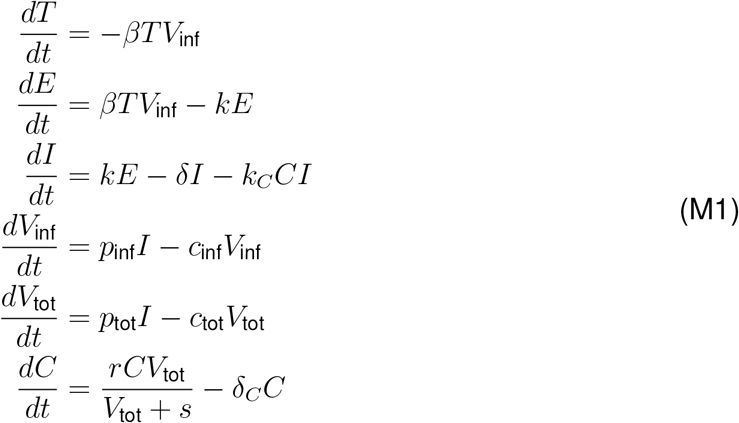

where *s* determines the viral load at which effector cell growth becomes virus independent and *r* is the maximal growth rate of effector cells [25].

### Parameter inference and model selection

An Approximate Bayesian Computation Sequential Monte Carlo (ABC-SMC) algorithm is used to infer model parameters from the human challenge data [26, 27]. This method is an extension of the original ABC algorithm and uses a sequence of decreasing tolerances to obtain intermediate distributions that ultimately converge on the target posterior distribution. Parameter sets are sampled from the intermediate distribution of the previous generation and perturbed using a multivariate normal kernel, the covariance matrix of which is selected here to be the optimal local covariance matrix introduced in [28]. Choosing a suitable sequence of tolerances is important since tolerances that decrease too slowly will result in slower convergence, whilst a sequence that decreases too quickly will lead to lower acceptance rates. To avoid manually setting the tolerances beforehand, an adaptive approach is implemented where the tolerance for the current generation is equal to the 0.25 quantile of the distances from the previous generation [26]. For the first generation, the tolerance is equal to the *N* ^th^ smallest distance from 5*N* iterations of an ABC rejection sampling algorithm, where *N* = 5 × 10^3^ is the desired sample size [29].

Solutions of the mathematical model (MM) for infectious virus and total virus are compared to the measurements of viral load from the human challenge (HC) nasal samples using the following distance metric:

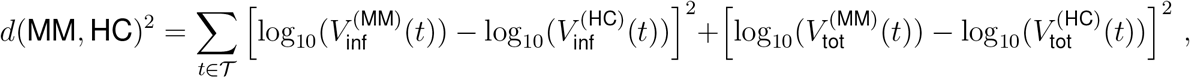

where 𝒯 is the set of times at which samples were taken. At times when the model solution falls below the limit of quantification (6 × 10^2^ RNA copies/mL and 5 PFU/mL), the solution is instead set equal to this limit. In doing so, no penalty is incurred when solutions drop far below the limit of quantification.

Given that the human challenge study collected samples from the first 14 days of infection, there is unlikely to be a significant decline in the adaptive immune response, therefore *δ*_*C*_ = 0. For the length of the eclipse phase, *in vitro* studies have showed that virus is released from infected cells 8 hours after infection, suggesting that it is suitable to fix *k* = 3 day^*−*1^ [30]. The viral load at which effector cell growth becomes independent of virus also remains constant at *s* = 10^4^ RNA copies/mL. For the remaining parameters, prior distributions are chosen as follows:

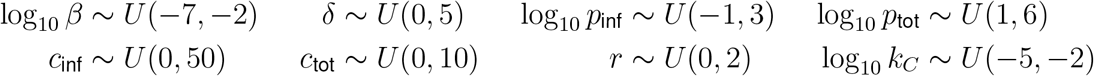

Previous studies have noted that only the product of *T* (0) and the viral production rate is identifiable from measurements of viral load [30]. Since it is more common to estimate viral production rates, the initial number of target and effector cells is set to *T* (0) = 10^6^ cells/mL and *C*(0) = 1 cell/mL respectively, whilst all other cell populations are initially zero. Local sensitivity analysis indicated that *V*_tot_(0) did not have a large impact on the viral load, likely because *V*_tot_ is not directly involved in the infection of target cells, and is therefore fixed at *V*_tot_(0) = 10^2^ RNA copies/mL. Participants of the human challenge study received a challenge dose of 10 TCID_50_ via intranasal drops, which approximately corresponds to 275 PFU/mL. However, it is unknown whether this level of virus reaches cells in the mid-turbinate. For this reason, the initial condition *V*_inf_(0) is also inferred with prior distribution *V*_inf_(0) ∼ *U* (−2, 2). The inference has been repeated using different prior distributions for *V*_inf_(0), but this did not result in significantly different approximate posterior distributions for the remaining model parameters (see Figure S1).

The ABC-SMC algorithm is used to infer parameters for each individual separately and therefore produces 16 separate approximate posterior distributions. To obtain a single distribution that is representative of all participants, a mixture distribution is constructed. Suppose that *f* (***θ***) is the probability density function for the true posterior distribution, where ***θ*** is the vector of model parameters. The density function for the mixture distribution is then given by:

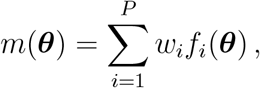

where *P* = 16 is the number of individuals that developed infection and *w*_*i*_ are the weights for each individual. Here, *w*_*i*_ = 1*/P* for all *i* since there is no evidence to suggest that any single participant is preferred over the others. Approximate samples from the mixture distribution can be obtained by first sampling an individual and then sampling from the corresponding approximate posterior sample.

To compare the mathematical model in M1 to an equivalent model without an immune response, an ABC-SMC model selection algorithm is used [27]. Model M2 is defined using the same system as M1 but removes the final ODE describing the effector cell population and therefore also ignores the clearance of infected cells by effector cells. The model selection algorithm follows the same procedure as the ABC-SMC algorithm for parameter inference but introduces a step prior to sampling candidate parameter sets where a candidate model is sampled. At the end of each generation, the Bayes factor for model M1 in favour of model M2 is calculated as follows:

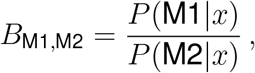

where the prior probabilities for model M1 and M2 are assumed to be equal. For each participant, the sequence of tolerances used in the model selection algorithm is the same as that obtained from the inference using model M1. A Bayes factor greater than 3 indicates positive evidence, whilst a value greater than 20 indicates strong evidence in favour of model M1 [27].

## Results

Measurements of viral load were obtained from the human challenge study described in [31]. In this study, 36 healthy, young participants were inoculated intranasally with a wild-type SARS-CoV-2 virus following a screening process that ruled out previous SARS-CoV-2 infection and recent respiratory infections. Two of these participants seroconverted between screening and inoculation and were removed from the study. Of the 34 individuals that continued in the study, 18 developed an infection. Six of these individuals were treated with remdesivir once two consecutive nose or throat swabs showed quantifiable SARS-CoV-2 detection by PCR, however, their viral loads were comparable to those of untreated participants and therefore no distinction was made between the treated and untreated participants in previous analyses [31]. Here, we continue to analyse the treated and untreated participants as a single cohort. Due to concentrations of viral RNA in the mid-turbinate remaining below the limit of quantification until day 6 and day 8, a further two participants are excluded since the model is not able to explain such long delays before viral growth. Although both nasal and throat swabs were collected, only measurements from the nasal swabs are used to parameterise our model.

The Bayesian inference provides us with a quantitative description of the in-host dynamics of SARS-CoV-2 virus. Model predictions for the infectious and total viral loads for 16 participants of the human challenge study are provided in Figure 1. From the posterior mixture distribution (Figure 2), we estimate that during the early stages of infection, productively infected cells are lost at an average rate of 1.17 day^*−*1^. By day 14, once an adaptive immune response is more established, this rate increases to 16.12 day^*−*1^. Individual infected cells produce infectious virus at rate 3.74 PFU/ml day^*−*1^ which is considerably lower than the production of viral RNA at rate 2.57 × 10^3^ RNA copies/mL day^*−*1^. The within-host basic reproduction number represents the number of secondary cellular infections a single infected cells causes in a population of fully susceptible cells, and is given here by *R*_0_ = *βT* (0)*p*_inf_*/δc*_inf_. Using the mixture distribution to derive an estimate of the within-host basic reproduction number gives *R*_0_ = 11. Note that this estimate will likely decrease as the infection progresses due to the shortened lifespan of productively infected cells.

**Figure 1:**
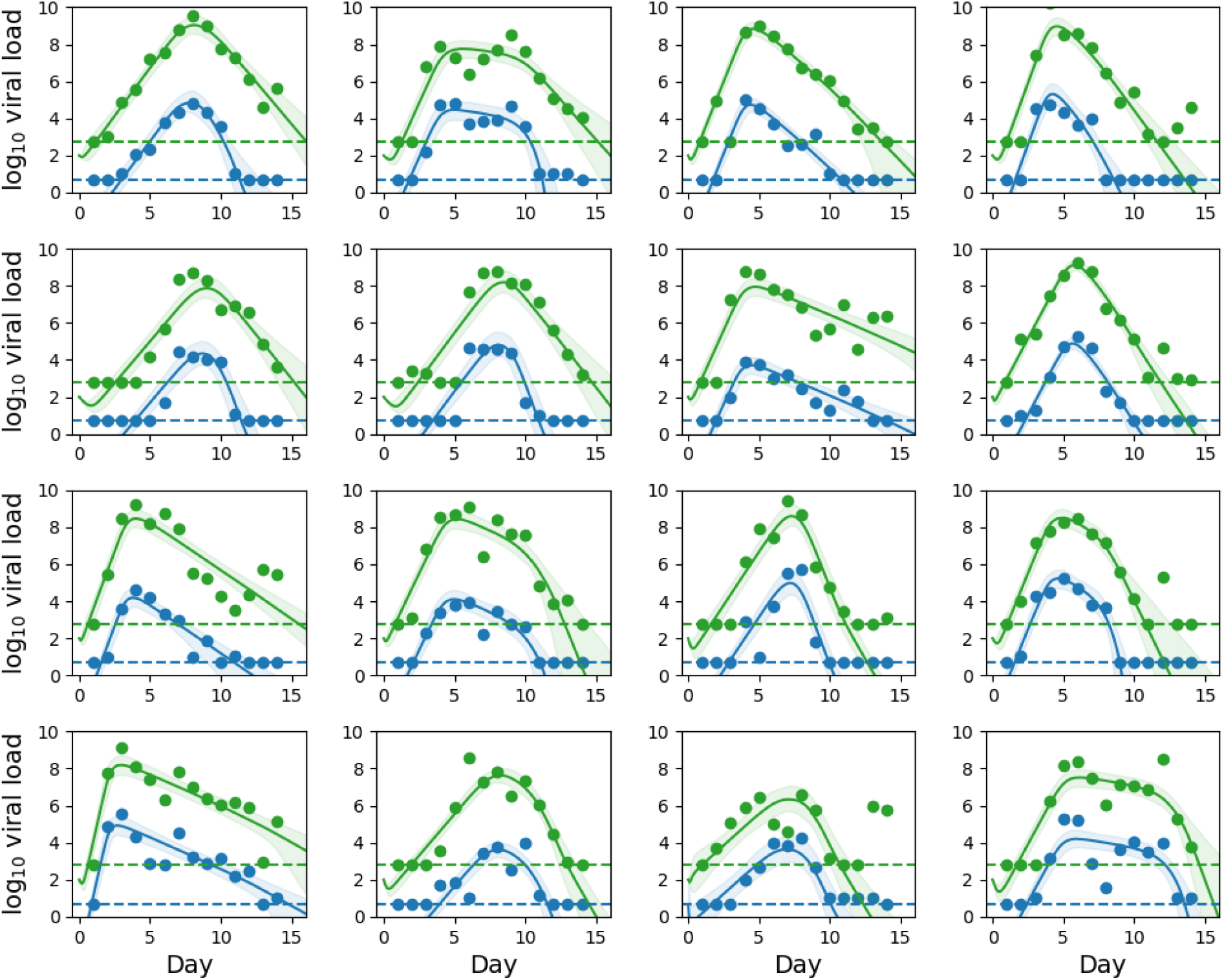
Posterior predictions: Predicted viral dynamics of infectious virus (blue) and total virus (green) for participants of the human challenge study, constructed using approximate posterior samples from the ABC-SMC. Solid lines represent pointwise median predictions and shaded regions indicate 95% credible regions. Horizontal dashed lines indicate the limit of quantification.

**Figure 2:**
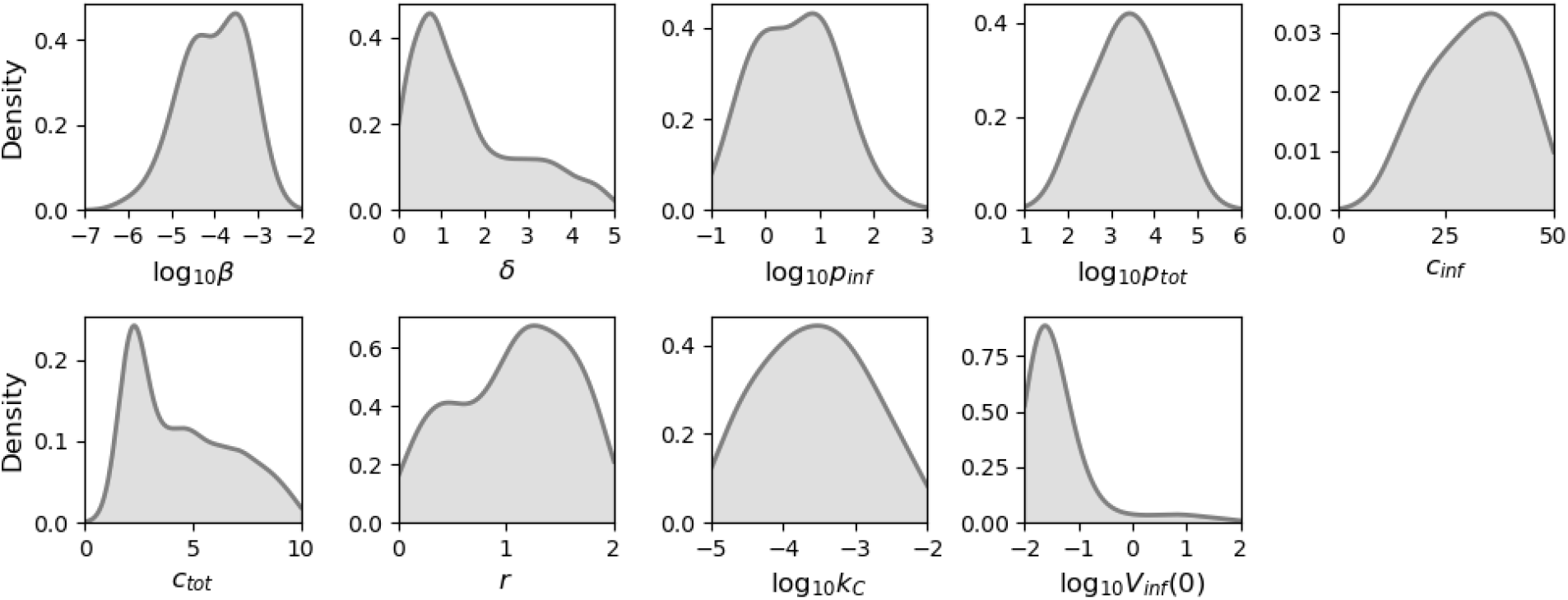
Inferred parameter distributions for human SARS-CoV-2 infection: Approximate posterior distributions for model parameters inferred using an ABC-SMC algorithm from midturbinate viral loads of human challenge study participants.

For many participants, the mathematical model predicts that infectious and total viral loads follow an initial exponential growth phase followed by an exponential decay phase. Since this behaviour can be captured by a model that excludes immune responses, a model selection algorithm was used to determine if the inclusion of effector cells was necessary (see Methods). For six individuals, there was very strong evidence in favour of the model with the immune response, whilst for a further individual there was positive evidence (see Figure S2). For the remaining nine participants, there was not sufficient evidence to suggest that either model was preferable over the other. As a result of this, we believe that the inclusion of an adaptive immune response is appropriate.

Predictions from the mathematical model can also be used to estimate timescales of infection. Here, we define the time that an individual first tests positive by a PCR test as the time at which total virus (*V*_tot_) first exceeds the limit of quantification. Similarly, the first time a sample is culture positive is assumed to be equivalent to the first time the level of infectious virus (*V*_inf_) exceeds the limit of quantification, and can be thought of as the earliest time an individual is capable of transmitting the virus. The duration that an individual remains PCR (culture) positive is then defined as the number of days the total (infectious) viral load continuously exceeds the limit of quantification. Estimates of these timescales are provided in Figure 3, along with an estimate of the incubation period, defined as the time to reach peak RNA copies/mL. Individuals are first PCR positive 1.1 days (interquartile range: 0.8–1.7 days) after infection and become infectious after 2.1 days (1.7–3 days). The incubation period lasts for 5.7 days (4.6–7.8 days) and is therefore consistent with the idea of pre-symptomatic transmission. Individuals remain infectious for 8.3 days (7.5–9.6 days) but would continue to test positive using a PCR test for 12.4 days (10.6–14.3 days). Some individuals remain PCR positive for up to 27 days following infection.

**Figure 3:**
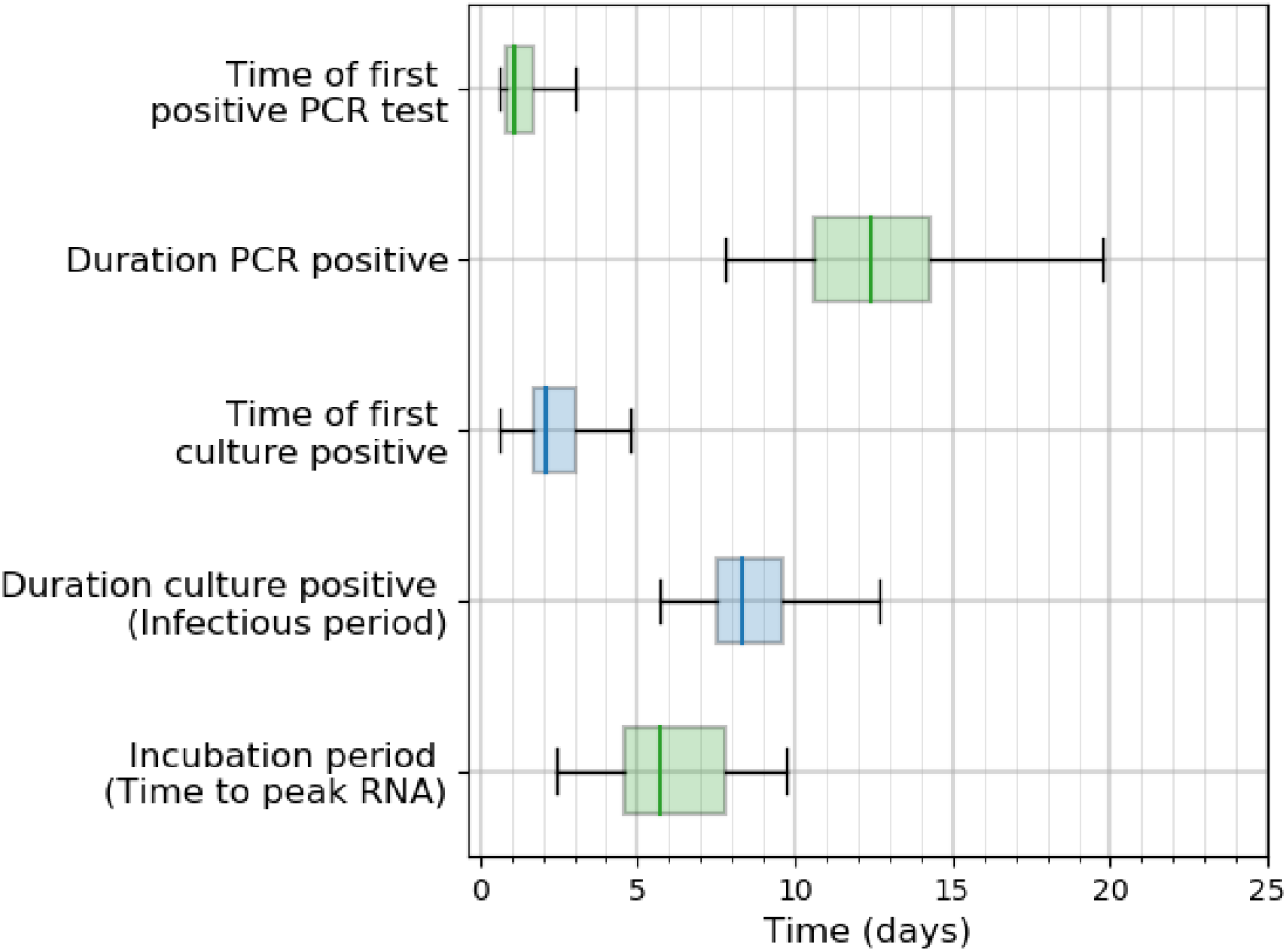
Timescales of infection: Estimates for the time of first positive test and the duration an individual remains positive. The timescales are derived from model predictions of both total (green) and infectious (blue) virus.

Whilst human challenge studies have many advantages, the strict screening process results in a relatively homogeneous population and so predictions from our model may not be representative of a more general population. For this reason, model predictions are validated against the Assessment of Transmission and Contagiousness of COVID-19 in Contacts (ATACCC) study, a longitudinal cohort study of community contacts of SARS-CoV-2 cases [32]. Whilst this study spanned multiple waves to capture the viral dynamics of unvaccinated and vaccinated individuals infected with different variants of SARS-CoV-2, we only consider those individuals who are unvaccinated and infected with a pre-alpha variant, since these will be the most similar to participants of the human challenge study. To determine if our mathematical model can describe the viral dynamics of ATACCC participants, a brute force approach is used that initially involves sampling 10^4^ parameter sets from the mixture distribution provided in Figure 2, in order to evaluate the model. Since participants are first contacted following symptom onset of the index case, the exact date of infection is unknown. Therefore, delays of up to 10 days between infection and enrolment are introduced to the model solutions in increments of 0.1 days. Solutions for log10(*V*_tot_) are then compared to log-RNA copies/mL from URT samples using the Euclidean distance, where only pairs of parameter sets and delays that yield the lowest 1% of distances are retained. These retained values are used to create the predictions in Figure 4. For the majority of ATACCC participants, the model and its existing parameterisation is able to explain the data well, suggesting that results derived from the human challenge study may be applicable to a general population. Furthermore, the median delay between infection and enrolment is 5.4 days which is plausible given that in most individuals we only observe the decay in the viral load and this decay begins around the time of symptom onset.

**Figure 4:**
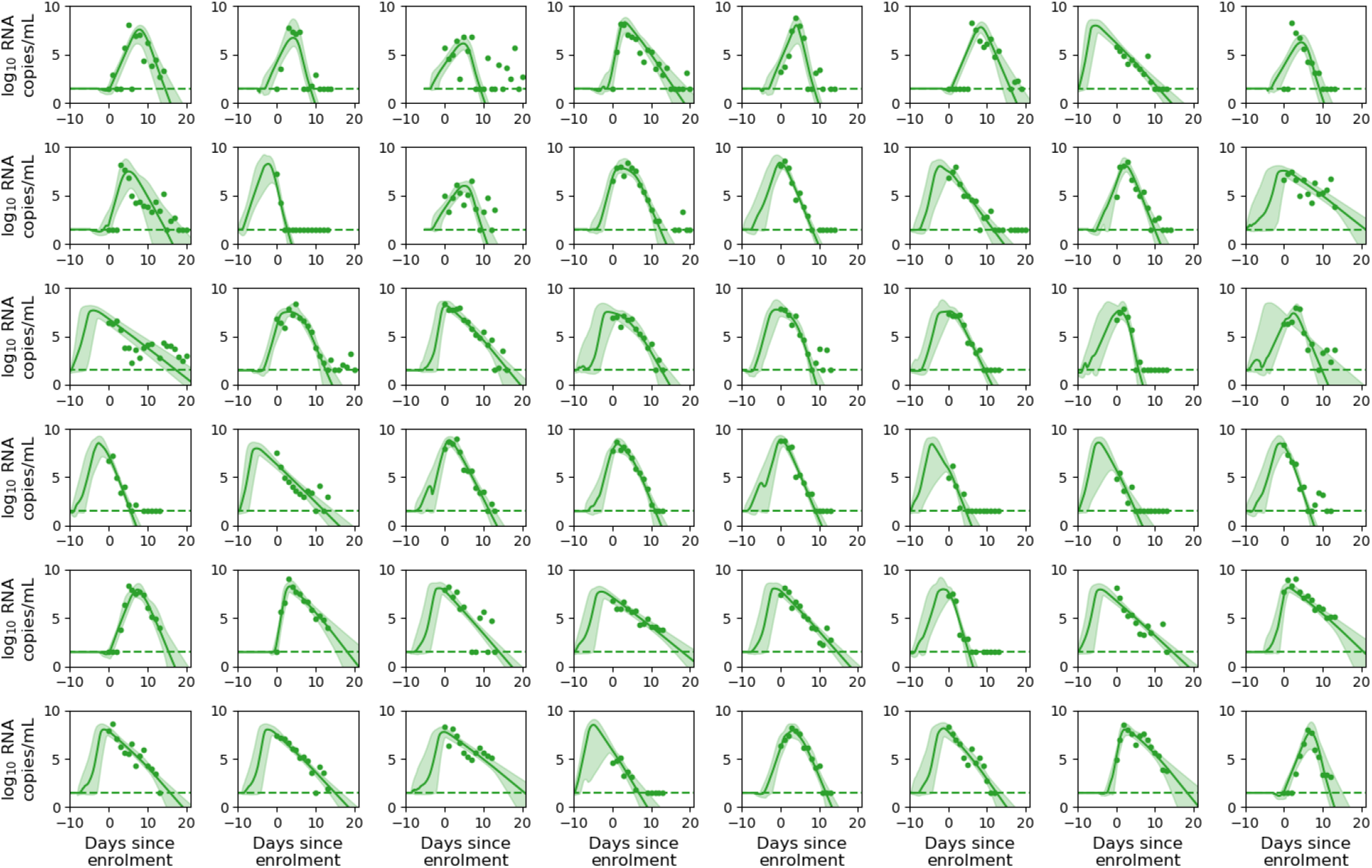
ATACCC validation: Predicted URT viral loads for participants in the ATACCC study. Predictions are constructed using samples from the mixture distribution presented in Figure 2 and allowing for a delay between infection and enrolment of up to 10 days. Solid lines represent point-wise median predictions and shaded regions indicate 95% credible regions. Horizontal dashed lines indicate the limit of detection.

## Discussion

In this paper, we have used viral load measurements from a human challenge study to parameterise a mathematical model of SARS-CoV-2 infection. Whilst existing models have distinguished between infectious and non-infectious virus, to the best of our knowledge this is the first modelling study that makes use of infectious virus titres during parameter inference. Our estimate of the within-host basic reproduction number (*R*_0_=11) is similar to estimates from previous modelling studies (8.5–14.2), as is our estimate of 0.85 days for the lifespan of productively infected cells during the early stages of infection (0.53–1.66 days) [11, 17, 33].

A review of existing studies determined that the median duration of wild-type SARS-CoV-2 virus detection in the URT is 14.5 days following symptom onset [19]. Whilst this estimate falls within the range of our PCR-positive distribution, it is based off studies from early on in the pandemic with small sample sizes and is therefore unlikely to be robust. A more comparable study involved measuring the viral RNA trajectories of basketball players and estimated that acute infection lasted for 11.2 days in asymptomatic cases and 14.3 days in symptomatic cases, both of which are consistent with our predictions [34]. One limitation of our current analysis is that in some participants, RNA copies/mL remained high after the end of the 14-day study period. As a result, there is greater uncertainty in our predictions after this time which leads to greater uncertainty in the tail of the PCR positive distribution. In order to obtain accurate estimates for the duration of infection, regular sampling must be extended past 14 days.

Previous estimates of an individual’s infectiousness have been based on the ability to successfully culture virus samples, however, such studies identify the presence of replicative competent virus and do not quantify the viral load [14]. Infectious virus may therefore be detectable but not at sufficient levels for someone to be infectious. As a result, estimates of the infectious period based on these studies are generally longer than that predicted here [14, 35]. A recent within-host model defines the infectious period to be the time when an individual’s transmission probability exceeds 10% of its maximum value, obtaining estimates ranging between 1.9 days and 7.9 days that lie towards the lower end of our predicted distribution [18]. Ultimately, the duration of the infectious period depends on how infectiousness is defined. Approaches based on viral load are preferable because they can be used to represent infectiousness on a continuous scale (such as a probability), whereas culture positivity only provides a binary outcome. Furthermore, viral load curves for infectious virus are beneficial as they provide a measure of the amount of virus capable of causing infection. Therefore, whilst our assumption that an individual is infectious if their infectious viral load exceeds 5 PFU/mL may be too simple, the predictions of infectious virus obtained here can be used alongside existing dose-response methods to provide more accurate estimates of infectiousness [8, 36].

Future work can use infectiousness curves as a link between within-host and population dynamics to develop more realistic multi-scale models of SARS-CoV-2 [9, 37]. With further information regarding T-cell responses and antibody levels following vaccination, such multi-scale models could be extended to consider the impact of waning immunity on susceptibility and transmission [38]. The results presented here are specific to wild-type SARS-CoV-2 virus, however, there is wide interest in variants of the virus since these have been associated with increased transmission and can rapidly spread throughout a population. One advantage of mechanistic models is that the parameters have simple interpretations, therefore, when the Delta variant reportedly showed a 10-fold increase in spike-mediated entry compared to a wild-type strain, this can be explicitly accounted for in the cellular infection rate [39]. The viral dynamics framework described here has also been extended to account for the genetic drift of viruses caused by mutations that occur during replication [40]. Applying this to SARS-CoV-2 virus may help understand the potential for new variants to arise within-host, with a recent report suggesting that the spike protein evolves to resist antibodies in immunocompromised individuals that have persistent infections [41]. Altogether, within-host models offer important insights into the interaction between SARS-CoV-2 virus and the host, however, it is the subsequent applications of these models that are most relevant for informing policy and decision making.

## Data Availability

All data produced in the present study are available upon reasonable request to the authors

