## Supplementary Figures for "A within-host model of SARS-CoV-2 infection"

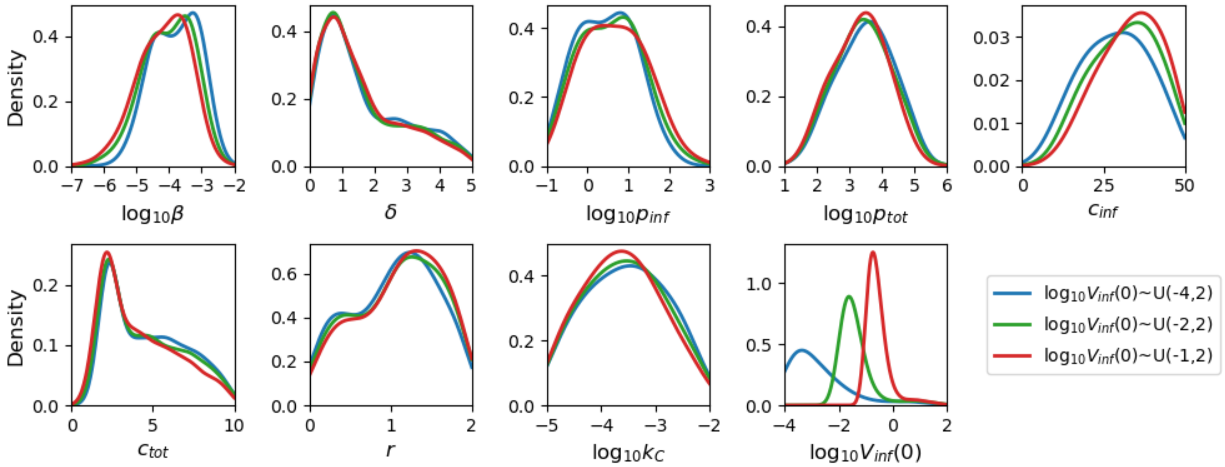


**Figure S1: Comparison of $V_{\mathbf{inf}}(0)$ prior distributions**: Approximate posterior distributions for model parameters inferred using an ABC-SMC algorithm with different prior distributions for the initial concentration of infectious virus.

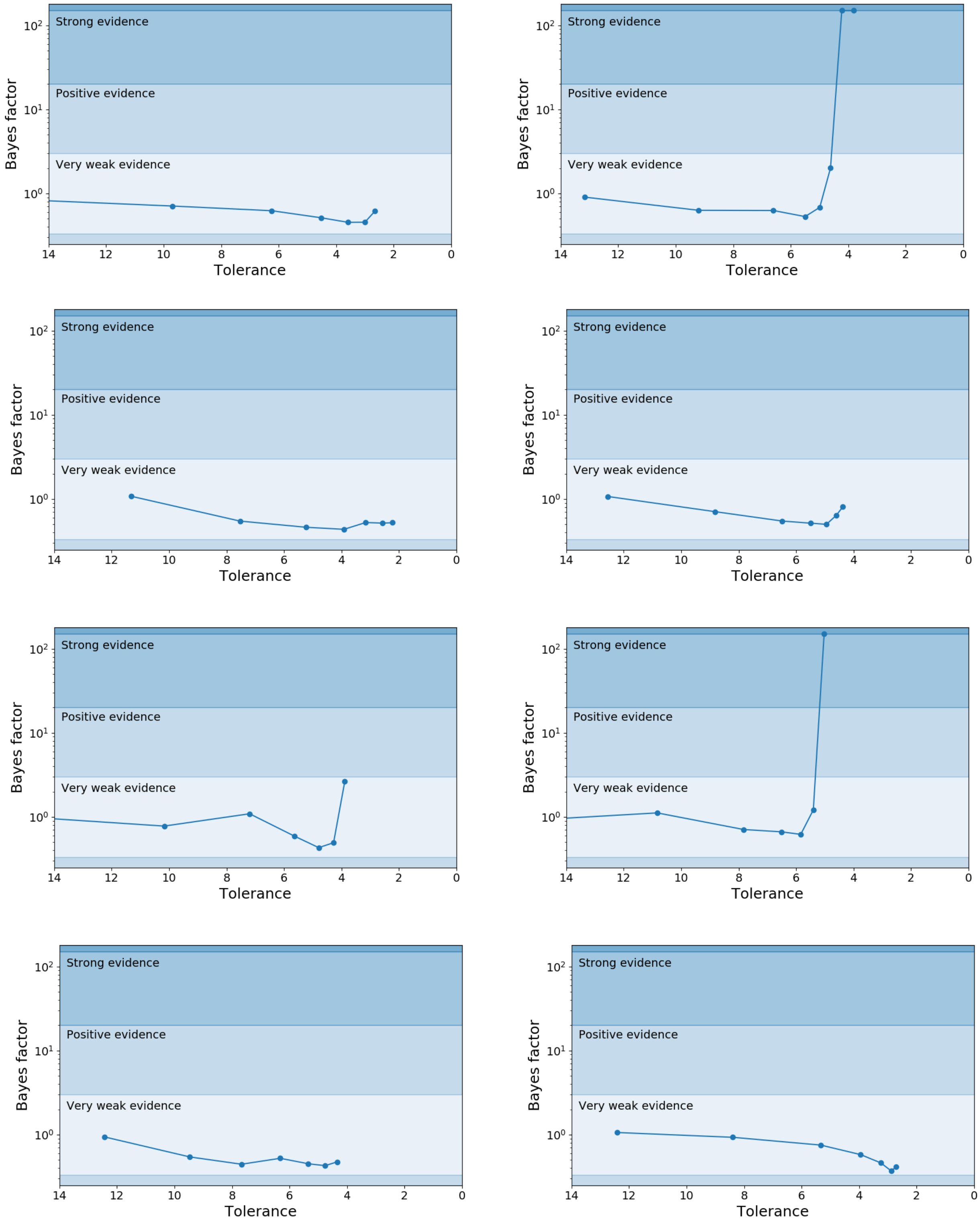


**Figure S2a: ABC-SMC model selection**: The Bayes factor is plotted against the tolerance for each generation of the ABC-SMC model selection algorithm. The strength of the evidence in favour of a model with an immune response is also indicated. For each participant, the sequence of tolerances is the same as that obtained from the ABC-SMC parameter inference.

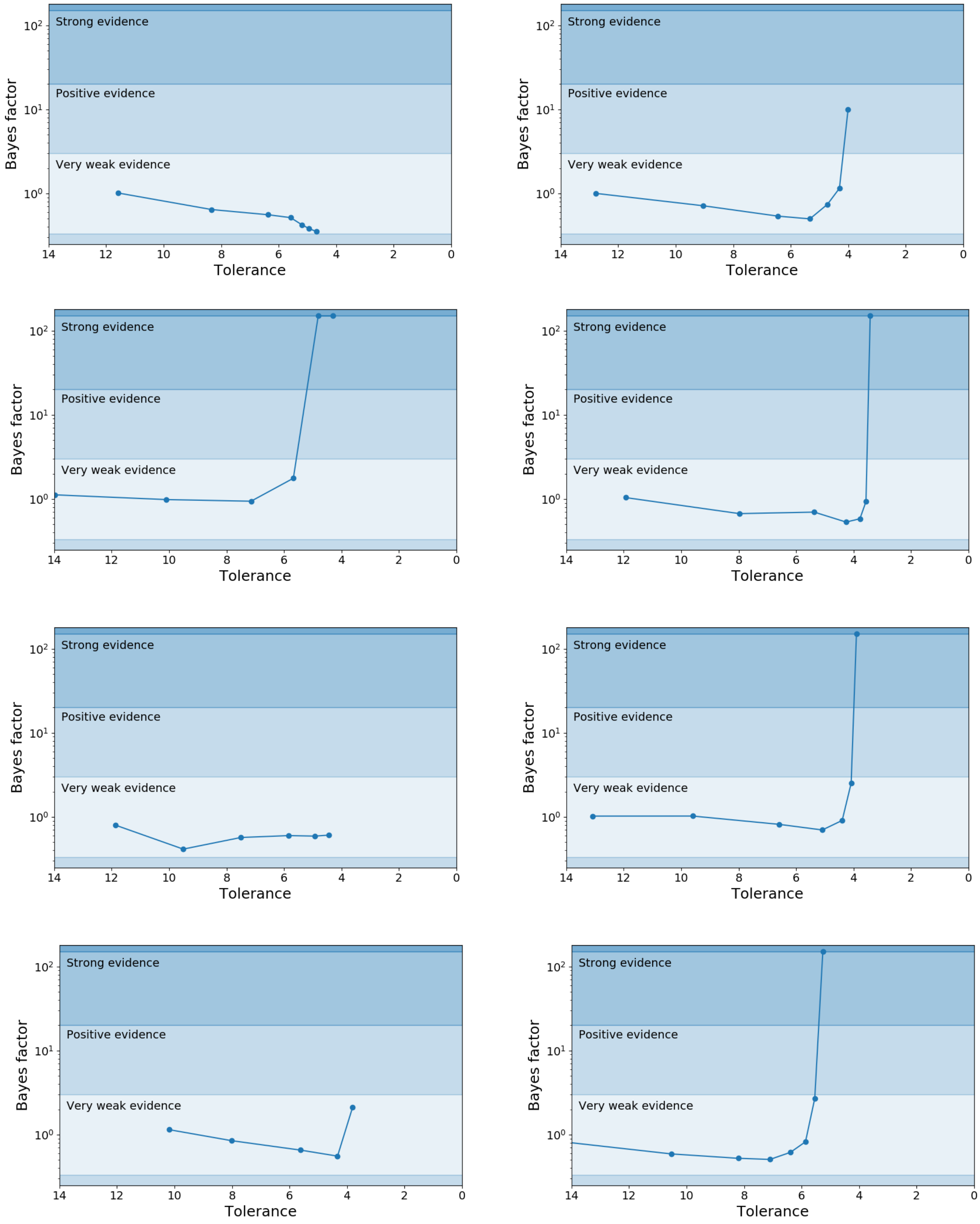


**Figure S2b: ABC-SMC model selection**: The Bayes factor is plotted against the tolerance for each generation of the ABC-SMC model selection algorithm. The strength of the evidence in favour of a model with an immune response is also indicated. For each participant, the sequence of tolerances is the same as that obtained from the ABC-SMC parameter inference.
